# Prevalence and predictors of tuberculosis in adults and adolescents with sputum trace Ultra results in two high-burden clinical settings

**DOI:** 10.1101/2025.05.16.25327787

**Authors:** Caitlin Visek, Ronit R. Dalmat, Annet Nalutaaya, Kamoga Caleb Erisa, Patrick Biché, Gabrielle Stein, Amanda Ganguloo, Robin Draper, Mariam Nantale, Adrienne E. Shapiro, Douglas Wilson, Achilles Katamba, Paul K. Drain, Emily A. Kendall

**Affiliations:** Division of Infectious Diseases, Department of Medicine, Johns Hopkins University School of Medicine, Baltimore, Maryland, USA; Department of Global Health, University of Washington, Seattle, Washington, USA; Uganda Tuberculosis Implementation Research Consortium, Walimu, Kampala, Uganda; Department of Epidemiology, Johns Hopkins Bloomberg School of Public Health, Baltimore, Maryland, USA; Umkhuseli Innovation and Research Management, Pietermaritzburg, South Africa; Department of Internal Medicine, Harry Gwala Regional Hospital, University of KwaZulu-Natal, Pietermaritzburg, South Africa; Clinical Epidemiology & Biostatistics Unit, Department of Medicine, Makerere University College of Health Sciences, Kampala, Uganda; Department of Epidemiology, University of Washington, Seattle, Washington, USA

## Abstract

**Background:** Some patients who test trace-positive on Xpert MTB/RIF Ultra (“Ultra”), a highly sensitive molecular diagnostic platform, may not have tuberculosis (TB) disease. A better understanding of the prevalence of TB disease, associated clinical characteristics, and utility of additional diagnostic tests among people with trace sputum (PWTS) could aid clinical decision-making.

**Methods:** We enrolled adults and adolescents with trace-positive sputum on initial TB diagnostic evaluation in Uganda and South Africa. Participants were extensively evaluated at enrollment; those with uncertain TB status were followed off treatment, with interval reevaluations by TB clinicians, for up to three months. We assessed TB prevalence and associated patient characteristics and diagnostic results.

**Results:** Among 311 PWTS, TB was identified by sputum culture at enrollment in 20% of participants (61/311, 95% CI 15-24%). Within three months, 48% (145/301, 95% CI 43-54%) had been judged by clinicians to warrant TB treatment, and among those followed until microbiologic outcomes, 30% (68/227, 95% CI 24-36%) had positive culture and 41% (99/240, 95% CI 35-47%) had positive culture or Ultra. Having TB symptoms, advanced HIV, and no recent TB history were associated with microbiologically-confirmed TB disease, as were an abnormal chest x-ray (in those without recent TB) or elevated CRP.

**Conclusions:** Roughly half of PWTS were started on TB therapy. Given the low observed mortality rate, some low-risk people with negative results on widely available diagnostic tests could safely defer treatment. Multimodal testing, repeated evaluations, and longer follow-up duration are needed to fully assess the TB burden in PWTS.

**Key points:** Half of patients with trace results on initial Xpert MTB/RIF Ultra diagnostic testing were diagnosed with TB within three months. Negative HIV status, lack of prior TB treatment, negative chest x-ray, and low C-reactive protein identified patients at lower risk.

## Background

Tuberculosis (TB) detection remains a challenge globally, with an estimated 29% of cases never being formally diagnosed [1]. Expanding access to rapid molecular diagnostics is a World Health Organization (WHO) priority, as these tools allow for accurate diagnosis of TB, early detection of drug resistance, and prompt initiation of effective therapy [1]. Xpert MTB/Rif Ultra (“Ultra”; Cepheid), a widely used rapid diagnostic test, achieves sensitivity close to that of culture, partly through incorporation of multi-copy *Mycobacterium tuberculosis* (MTB) gene targets [2, 3]. The lowest level of MTB detection, a category called “trace,” is reported when Ultra detects only these multi-copy targets and not the single-copy-per-genome *rpoB* target.

Trace results have uncertain clinical significance, as the presence of MTB DNA may reflect very low disease burden or remnant material from non-viable bacilli [4]. In diagnostic evaluations of patients with presumptive pulmonary TB, the trace category has accounted for 8 to 41% of all positive Ultra results [5–8], and as many as 9 in 10 of these trace results have been associated with negative sputum cultures and considered false positives [5]. However, many of these studies were small and included limited diagnostic evaluations. More extensive diagnostic evaluation, as well as interval reevaluations prior to treatment, could clarify whether some trace results represent disease even when cultures are negative. In addition, while several studies have found that people previously treated for pulmonary TB are especially likely to have trace sputum results and negative cultures [6, 8, 9], little is known about other patient characteristics or test results that might alter the likelihood of TB disease in people with trace-positive sputum (PWTS). Given these gaps in knowledge, current WHO guidelines recommend treating all presumptive TB patients with trace results, with the narrow exception of HIV-negative adults treated for TB within the past five years [10]. Understanding when a ‘Trace’ result is most likely to indicate TB disease would allow for more nuanced treatment decisions.

Therefore, we conducted a prospective cohort study in two countries to better delineate the TB disease status of PWTS during routine TB diagnostic evaluations. Following their initial trace result, participants underwent a systematic diagnostic evaluation, and if their TB status remained uncertain, treatment was deferred, and they were reassessed for evidence of TB disease one month and three months later. We estimated the proportions of patients diagnosed with TB clinically or microbiologically within three months and identified patient characteristics and ancillary diagnostic test results associated with TB disease.

## Methods

### Study design, participants, and procedures

We recruited adult and adolescent PWTS from routine TB diagnostic evaluations at seven health facilities in Kampala, Uganda, from February 2022 to July 2024, and from outpatient clinics of Harry Gwala Regional Hospital and its catchment area near Pietermaritzburg, South Africa, from November 2022 to June 2024. Study members attempted to recruit and enroll all eligible PWTS. Eligible patients had not yet initiated TB treatment, did not require hospitalization, and had not received TB treatment (both sites) or preventive therapy (South Africa only) within the previous three months.

Within 14 days of the initial sputum Ultra result, consenting participants underwent an extensive baseline evaluation that included a standardized interview, physical exam, digital chest x-ray (CXR), repeat sputum Ultra testing, sputum cultures (liquid and solid culture on each of two expectorated specimens), *Mtb* immunoreactivity testing (QuantiFERON-TB Gold+; Uganda site only), chest CT (Uganda site only), HIV testing, and serum C-reactive protein (LumiraDx [South Africa], ichroma [Uganda]; CRP) measurement. Patients with HIV also underwent urine lipoarabinomannan (LAM) testing and CD4+ T-cell count measurement.

Participants who remained untreated after this baseline diagnostic evaluation (by choice or recommendation of a clinician) were reassessed at one and three months with interview, sputum Ultra, sputum culture, CRP, and (at three months) repeat CXR.

### Monitoring and treatment initiation

At the South African site, a three-person panel of study doctors evaluated participants at each visit and determined whether to initiate or continue to defer TB treatment. At the Ugandan site, participants remained under the care of TB clinicians at the referring health facilities, who could start treatment at any time but were encouraged to await study investigations when clinically appropriate, and three-person advisory panels of pulmonologists and a radiologist reviewed new study data and made additional treatment recommendations.

Although participants were enrolled to follow for a year or more, this analysis focuses on TB diagnosed within three months, a period interpreted as indicating “prevalent” disease (present at the time of the initial trace Ultra result).

### Outcomes and missing data

Primary analyses use a **composite** TB reference standard: having MTB complex on sputum culture, a recommendation to start TB treatment (on any microbiological and/or clinical basis), or death attributed to tuberculosis. Sensitivity analyses use more limited **microbiological** (positive if MTB complex grew on sputum culture or Ultra was positive at a level greater than trace) and **culture-only** reference standards. All reference standards are assessed at two timepoints: at baseline (considering all tests on specimens from, and treatment decisions based on, the baseline study visit) and within three months (cumulatively). Participants are excluded from any microbiological outcome for which results are missing or uninterpretable, from all three-month outcomes if they missed the three-month visit, and from microbiological outcomes at three months if they initiated treatment before the three-month visit without microbiological confirmation (**Supplemental Table 1).**

### Data analysis

Continuous variables are summarized as median and interquartile range (IQR). Prevalence estimates are reported with 95% Wald confidence intervals.

We evaluated patient characteristics associated with TB in the cohort using log-binomial and robust Poisson regression. We developed analogous models for each of the three TB reference standards, classifying by the culture-only reference at baseline (to avoid bias from earlier culture-negative TB diagnoses) and the composite and microbiological reference standards at three months. We used forward stepwise selection of patient characteristic variables for each reference standard separately, considering the following variables *a priori* for inclusion: age (dichotomized at 35 years), gender, country, history of tobacco smoking, recent TB (treated within the past five years), symptoms (cough, weight loss, night sweats, or fever at baseline), recent household TB contact (diagnosed within the past six months), HIV status, and CD4 count (modelled only as an interaction with HIV status and categorized as <200 cells/mm^3^, ≥200 cells/mm^3^, or unknown). All variables selected for any of the three reference standards were included in all final models. A small number of participants with missing covariate data (**Table 1**) were excluded, except in a sensitivity analysis using multiple imputation (‘mice’ package; **Supplement**).

**Table 1:** Patient characteristics and baseline diagnostic results for trace-positive participants

| Variable | Combined cohort (N=311) | Uganda (N=170, 55%) | South Africa (N=141, 45%) |
| --- | --- | --- | --- |
| Age, years (median [IQR]) | 37 [30, 47] | 35 [29, 43] | 40 [33, 53] |
| Male sex (N [%]) | 155 (50) | 98 (58) | 57 (40) |
| Symptoms present on initial screen |  |  |  |
| Cough (N [%]) | 243 (78) | 154 (91) | 89 (63) |
| Fever (N [%]) | 118 (38) | 71 (42) | 47 (37) |
| Night sweats (N [%]) | 88 (28) | 42 (25) | 46 (33) |
| Weight loss (N [%]) | 146 (47) | 99 (58) | 47 (33) |
| Underweight (BMI <18.5) (N [%]) | 52 (17%) | 37 (22%) | 15 (11%) |
| HIV positive (N [%]) | 178 (57) | 75 (44) | 103 (73) |
| CD4 count <200 cells/mm<sup>3</sup> (N [%]) | 35 (21) | 16 (24) | 19 (19) |
| CD4 count >200 cells/mm<sup>3</sup> (N [%]) | 133 (79) | 50 (76) | 83 (82) |
| Taking ART (N [%]) | 144 (86) | 57 (78) | 87 (92) |
| Pregnant (N [%]) | 11 (4) | 4 (2) | 7 (5) |
| Smoking status |  |  |  |
| Never (N [%]) | 203 (65) | 116 (68) | 87 (62) |
| Current (N [%]) | 59 (19) | 18 (11) | 41 (29) |
| Former (N [%]) | 49 (16) | 36 (21) | 13 (9) |
| Smoking history of 20 or more pack-years among current or former smokers (N [%]) | 69 (73) | 37 (69) | 32 (78) |
| Alcohol use: Positive AUDIT-C score<sup>1</sup> (N [%]) | 104 (33) | 55 (32) | 49 (35) |
| Prior TB treatment (N [%]) | 113 (36) | 64 (38) | 49 (35) |
| Prior TB treated within the last 5 years (N [%]) | 72 (24) | 49 (29) | 23 (17) |
| Recent household contact with TB (within the past 6 months) (N [%]) | 25 (8) | 13 (8) | 12 (9) |
| Abnormal baseline chest x-ray result (N [%]) | 196 (65) | 111 (68) | 85 (61) |
| Baseline laboratory evaluation |  |  |  |
| C-reactive protein >5mg/L (N [%]) | 155 (50) | 85 (50) | 70 (50) |
| Repeat Xpert Ultra result |  |  |  |
| Negative (N [%]) | 215 (71) | 117 (69) | 98 (74) |
| Trace positive (N [%]) | 35 (12) | 22 (13) | 13 (10) |
| Positive >trace (N [%]) | 52 (17) | 31 (18) | 21 (16) |
| Positive culture (N [%]) | 61 (20) | 41 (24) | 20 (14) |

Small number of participants were missing data for: underweight (3), CD4 count among participants with HIV (10), ART status among participants with HIV (10), pregnancy status among female participants (1), pack-year history among current or former smokers (13), history of prior TB (1), timing of prior TB treatment among participants with a history of TB (6), baseline CXR (9), C-reactive protein (1), repeat Xpert Ultra (9). Missing data was excluded from the denominators when calculating percentages.
BMI = Body mass index
AUDIT-C = Alcohol Use Disorders Identification Test
1. Positive AUDIT-C score defined as 4 or greater for men and 3 or greater for women.

To the resulting “patient characteristics” models, we subsequently added either CXR results (normal/abnormal, “CXR-added model”) or CRP results (normal/elevated using a threshold of 5 mg/L, “CRP added model”) as an additional covariate. The CXR-added model included an interaction between CXR result and recent TB. We generated receiver operating characteristic (ROC) curves for each model and calculated areas under the ROC curves (AUCs) using the ‘pROC’ package (version 1.18.5).

Finally, we used our CXR-added regression model to estimate the risk of TB among a simulated cohort of patients with a specified set of low-risk features – namely, negative HIV status and normal baseline CXR – but no history of TB. Results are reported as the median and IQR of estimated individual-level risks. See Supplemental Methods for further detail.

All analyses used R Statistical Software version 4.3.1 (R Core Team, 2023).

### Ethical approval statement

The study was approved by the University of KwaZulu-Natal Biomedical Research Ethics Committee, the Makerere University School of Public Health Research and Ethics Committee, and the institutional review boards at the University of Washington and Johns Hopkins University.

## Results

A total of 311 PWTS (170 in Uganda; 141 in South Africa) were enrolled by the analysis cutoff dates. Another 796 (59 in Uganda; 737 in South Africa) had trace results at participating facilities but were not enrolled, primarily due to distance or inability to make contact (**Figure 1, Supplemental Figure 1**). At one and three months, 190 and 162 participants, respectively, remained alive and untreated and completed follow-up visits. Mortality during this three-month period was 0.6% (2/311), with one of the deaths attributed to TB.

**Figure 1:**
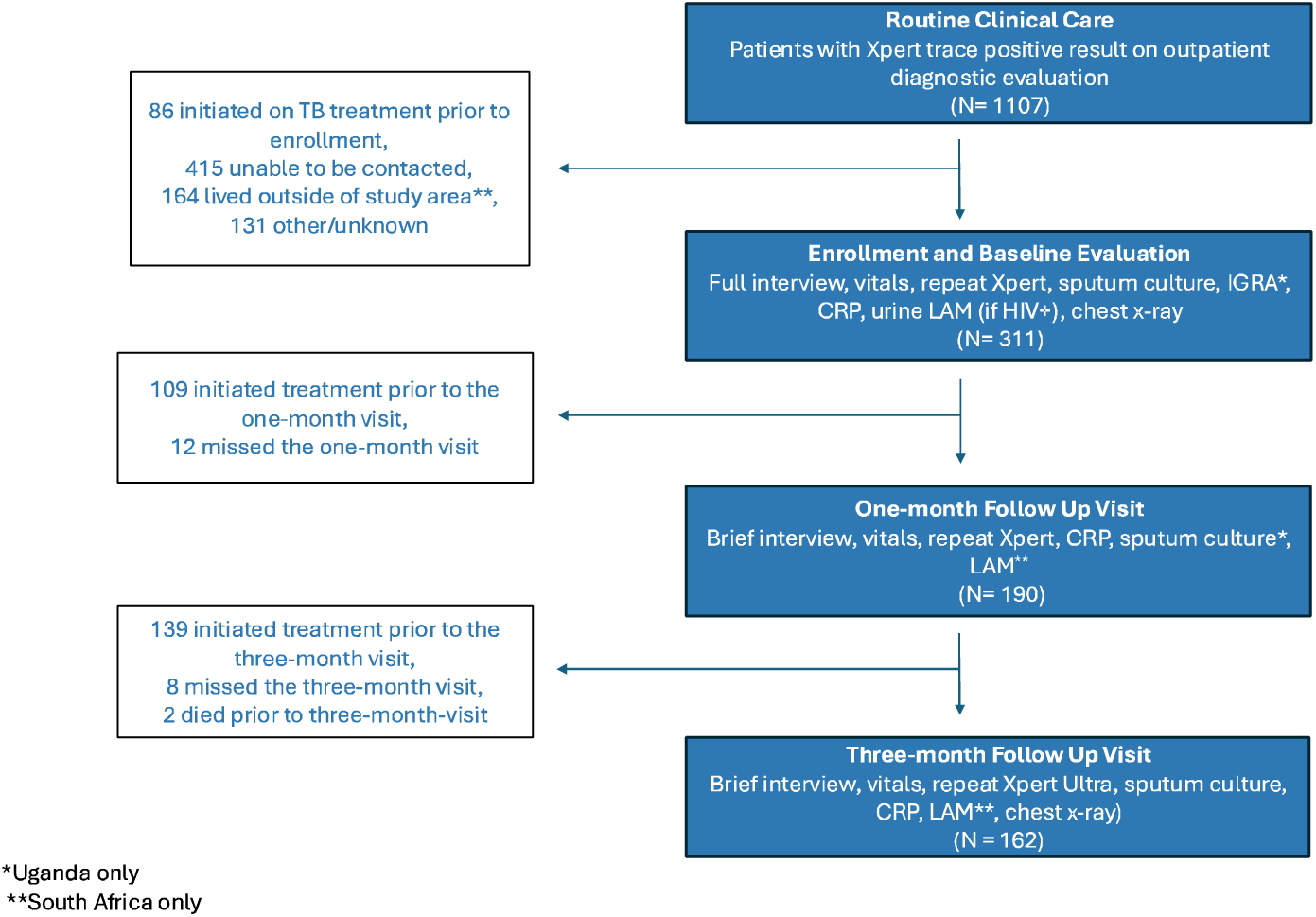
Study population overview. Flow diagram of patients with trace results on diagnostic evaluation, study enrollment, and visit completion, with reasons for exclusion

The cohort was 50% male (155/311) with a median age of 37 (**Table 1**). HIV prevalence was 57% (178/311), and most participants living with HIV were on antiretroviral therapy (ART) and had CD4 counts greater than 200 cells/mm^3^. Nearly a quarter (24%) had been treated for TB within the past five years, and most had one or more TB symptom. On repeat Ultra testing at baseline, most participants (215/302; 71%) tested negative, but 12% (35/302) tested trace positive (of whom 11 [31%] also had positive baseline sputum cultures) and 17% (52/302) had positive results at a level greater than trace (of whom 32 [62%] had positive cultures). Half of participants (50%; 156/310) had an elevated CRP, and 65% (196/302) had an abnormal CXR (**Table 1**).

Overall, 145 of 301 participants (48%, 95% CI 43-54%) were positive by the composite reference standard by three months (**Figure 2**). Twenty-five (17%) of these diagnoses were based on additional data obtained during follow up. Eighty-three (57%) were confirmed by Xpert (>trace) or culture (**Supplemental Figure 2**).

**Figure 2:**
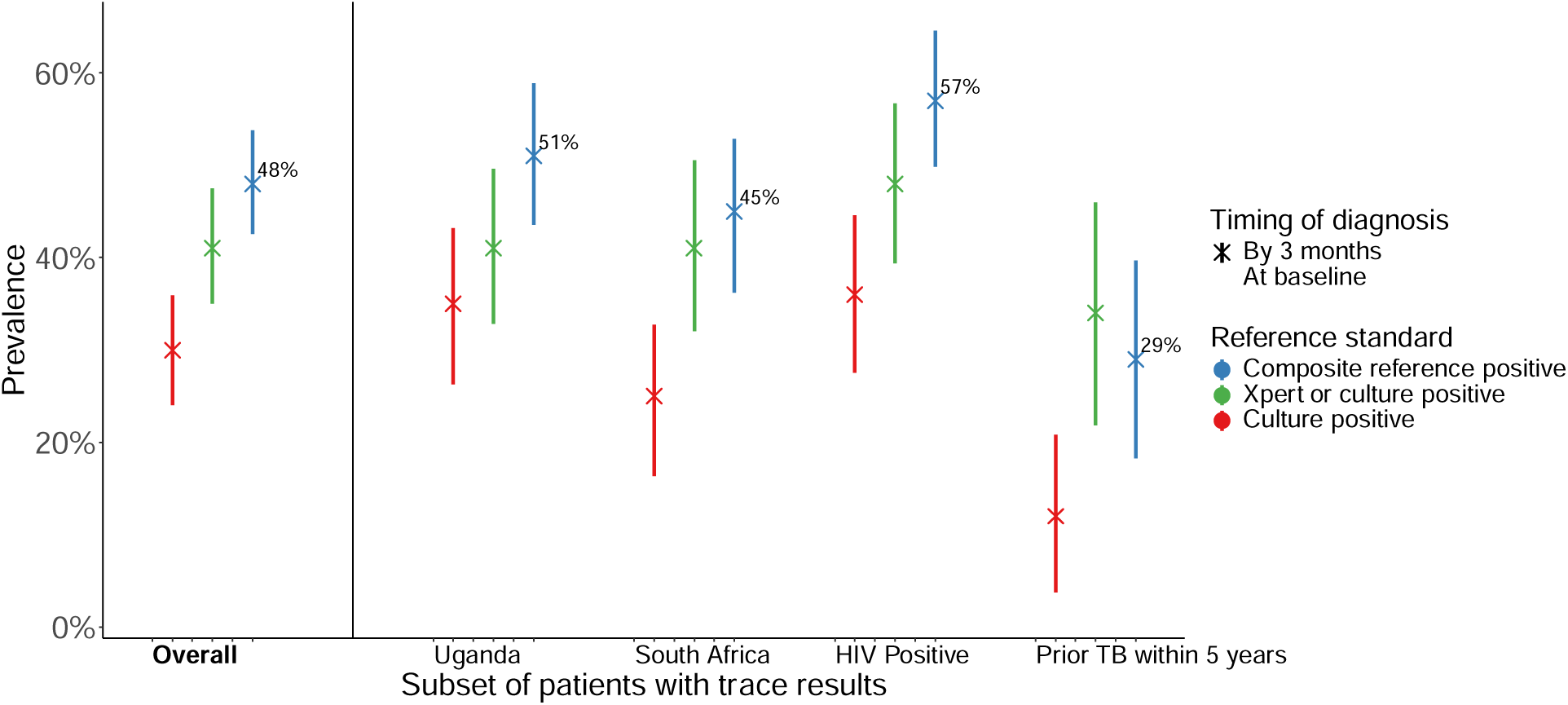
TB prevalence among patients with initial trace Xpert Ultra results, by multiple definitions. *TB prevalence by the composite, microbiological (Xpert or culture), and sputum-culture only reference standards assessed at baseline and by three months, for the overall cohort and several subgroups. Prevalence estimates are represented as a point estimate and plotted with lines showing their 95% confidence intervals. CD4-count-stratified and symptom-stratified results are shown in* ***Supplemental Figures 4-5***.

Of those who could be evaluated using the microbiological reference standard, TB prevalence was 26% (81/311, 95% CI 21-31%) at baseline and 41% (99/240, 95% CI 35-47%) by three months (**Figure 2**). Some patients with positive microbiological classifications (31/99 [31%] of those positive by 3 months) had positive (>Trace) Ultra results but negative cultures; 16 of these were considered falsely positive with no treatment recommended. When considering sputum culture only, TB prevalence was 20% (61/311, 95% CI 15-24%) at baseline. Among the entire cohort, 68/311 (22%) had at least one positive culture by three months, with seven participants having newly positive cultures during follow up; three-month prevalence by the culture reference standard (after excluding 84 who had started treatment without culture confirmation (72 participants) or were missing three-month culture results (12) was 30% (68/227, 95% CI 24-36%).

In univariate analysis, TB was associated with male gender, HIV positivity, low or unknown CD4 count, TB symptoms, and a lack of recent TB treatment history, as well as with diagnostic results of abnormal CXR (particularly in those without recent TB) and elevated CRP (**Table 2**, **Supplemental Tables 5-6**). A positive or trace repeat Ultra was associated with a prevalence ratio (PR) of having TB of 2.2 [95% CI 1.8-2.7] for the three-month composite reference standard and 5.9 (95% CI 3.6-9.6) by the baseline culture-positive reference standard (**Supplemental Table 3**).

**Table 2:** Multivariate log-binomial regression model coefficients for discriminating TB vs no TB, using composite reference standard over 3 months

| Covariate | Unadjusted prevalence ratio (95% CI), n=287<sup>3</sup> | Adjusted prevalence ratio (95% CI) for variables included in final model |  |  |
| --- | --- | --- | --- | --- |
|  |  | Patient characteristics model, n=295<sup>4</sup> | CXR-added model, n=288<sup>5</sup> | CRP-added model, n=294<sup>6</sup> |
| Uganda site | 1.2 (1.0, 1.6) | - | - | - |
| Age < 35 | 1.1 (0.9, 1.5) | 1.2 (0.8, 1.6) | 1.2 (0.8, 1.7) | 1.2 (0.8, 1.6) |
| Male gender | 1.3 (1.0, 1.7) | 1.3 (0.9, 1.8) | 1.2 (0.8, 1.7) | 1.2 (0.9, 1.7) |
| History of prior TB within 5 years | 0.5 (0.3, 0.8) | 0.6 (0.3, 0.9) | 0.9 (0.1, 2.9) | 0.6 (0.4, 0.9) |
| Presence of 1 or more TB symptom<sup>1</sup> | 3.9 (1.9, 10.4) | 3.1 (1.5, 7.8) | 3 (1.3, 8.5) | 2.7 (1.3, 7.1) |
| Recent household TB contact<sup>2</sup> | 0.7 (0.4, 1.2) | 0.8 (0.4, 1.5) | 0.9 (0.4, 1.7) | 0.8 (0.4, 1.5) |
| Any smoking history | 1.1 (0.9, 1.4) | - | - | - |
| HIV positive (all) | 1.6 (1.2, 2.1) | - | - | - |
| HIV positive with CD4 <200 cells/mm<sup>3</sup> | 2.5 (1.5, 4.0) | 2.0 (1.2, 3.3) | 1.9 (1.2, 3.2) | 1.7 (1.1, 2.9) |
| HIV positive with CD4 >200 cells/mm<sup>3</sup> | 1.3 (0.9, 2.0) | 1.4 (0.9, 2.1) | 1.4 (1, 2.2) | 1.5 (1.0, 2.2) |
| HIV positive with CD4 unknown | 2.5 (1.1, 5.1) | 2.0 (0.9, 4.1) | 2.3 (1.0, 4.7) | 2.0 (0.9, 4.2) |
| Abnormal CXR (overall) | 1.7 (1.3, 2.4) | - | - | - |
| Abnormal CXR in those without history of TB within 5 years | 2.1 (1.4, 3.2) | - | 1.8 (1.2, 2.8) | - |
| Abnormal CXR in those with history of TB within 5 years | 0.4 (0.1, 2.6) | - | 0.6 (0.1, 3.7) | - |
| CRP > 5mg/L | 2.0 (1.6, 2.7) | - | - | 1.7 (1.2, 2.5) |

1. Defined as cough, fever, weight loss, or night sweats at time of study enrollment
2. Defined as living with someone diagnosed with TB within the past 6 months
3. 24 observations dropped due to incomplete data
4. 16 observations dropped due to incomplete data
5. 23 observations dropped due to incomplete data
6. 17 observations dropped due to incomplete data

In multivariable regression, TB (defined by the composite reference standard by three months) remained positively associated with advanced HIV (CD4 <200 cells/mm^3^: PR 2.0 [95% CI 1.2-3.3]) and TB symptoms (PR 3.1 [95% CI 1.5-7.8]) and inversely associated with recent TB (PR 0.6 [95% CI 0.3-0.9]) (**Table 2**). When considering additional diagnostic test results, either an abnormal CXR (in those without recent TB: PR 1.8 [95% CI 1.2-2.8]) or an elevated CRP (PR 1.7 [95% CI 1.2-2.5]) was associated with greater likelihood of having TB. Results for alternative reference standards were largely similar, though associations with TB history and CXR results were weaker when using the microbiological reference standard that classified all people with positive Ultra results as having TB (**Supplemental Tables 5-6**). A sensitivity analysis using multiple imputation of missing data produced generally similar results (**Supplemental Table 7**).

The patient characteristics regression model classified TB status (composite reference standard, three months) with an AUC of 0.76 (95% CI 0.71-0.82) by. This AUC increased to 0.80 (95% CI 0.75-0.85) or 0.79 (95% CI 0.74-0.84) after adding CXR or CRP, respectively (**Figure 3**). AUCs were similar for the other reference standards (**Supplemental Figure 4)**.

**Figure 3:**
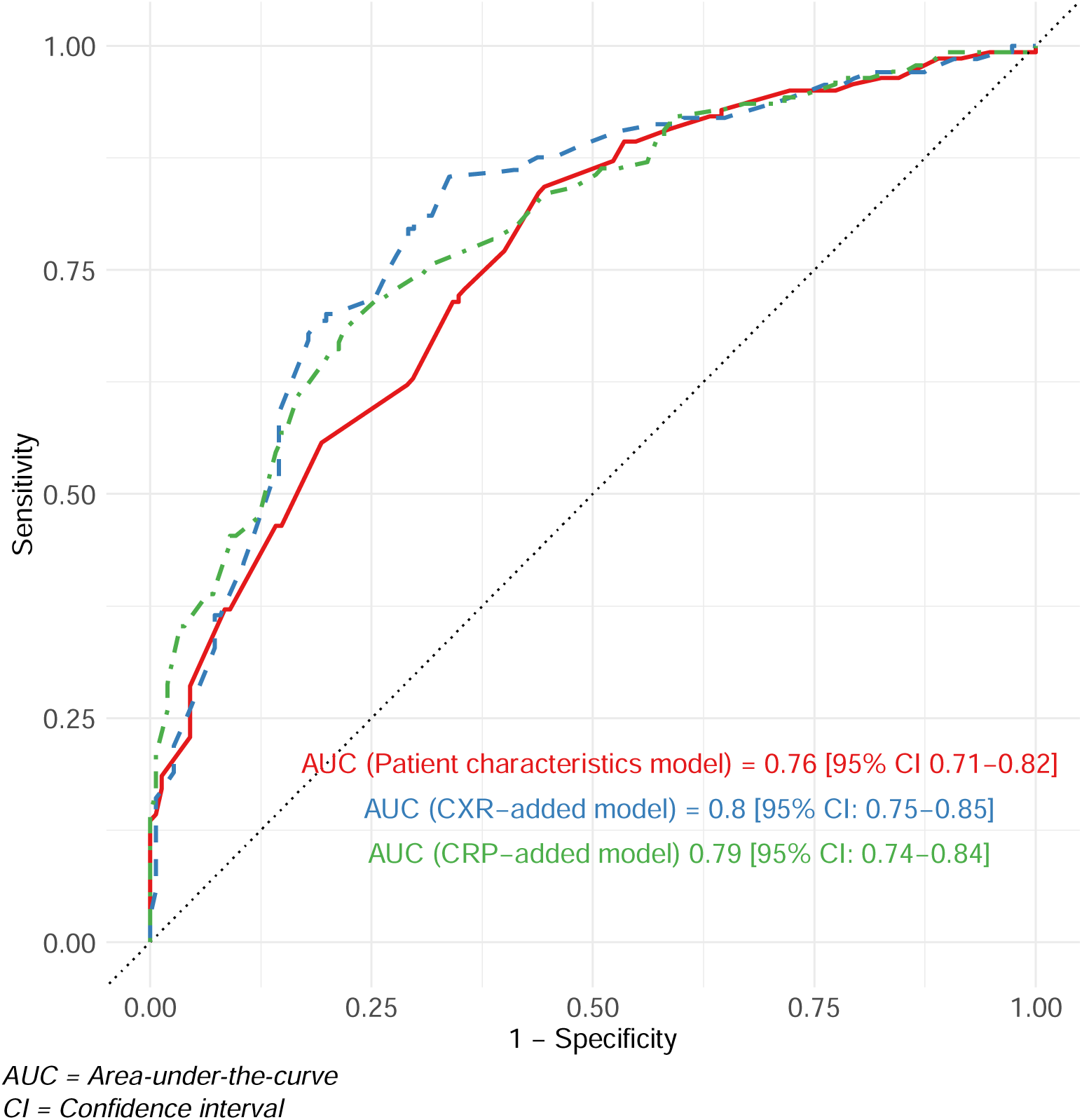
Receiver operating characteristic curves for the basic characteristics, CXR-added, and CRP-added models using the composite reference standard over 3 months

When we applied the CXR-added regression model to the simulated “low-risk” cohort of HIV-negative patients with normal baseline CXRs (but no recent history of TB), the median risk of TB (by the composite reference standard) was 27% (IQR 22-30%).

## Discussion

Our study offers a detailed look into the disease status and clinical and microbiological profiles of the largest cohort to date of patients with trace Ultra sputum diagnostic test results. More than 300 patients from multiple facilities in two high-prevalence countries were followed untreated for up to three months to clarify their TB status. Nearly half were diagnosed with TB within three months, including 41% with some microbiological confirmation. Baseline sputum culture (on which 20% of patients tested positive) did not capture the full burden of TB. Still, half of patients did not have sufficient evidence to diagnose TB despite three months of extensive testing. Across multiple TB definitions, patient characteristics associated with TB in this population included advanced HIV, TB symptoms at the time of enrollment, and having no recent history of TB treatment. Results of CXR and CRP testing further aided in risk-stratifying patients.

Accurately defining TB disease is a challenge when studying TB prevalence. Sputum culture, while often the gold standard, can be insufficiently sensitive to detect active pulmonary disease with low bacillary burden [11]. TB disease burden may also wax and wane [12, 13], limiting detection through one-time evaluations. We considered TB diagnosed within three months of a trace result, a timeframe chosen to allow for multiple reassessment opportunities and sufficient disease progression in cases of recent primary exposure, yet short enough that any TB that was identified was likely to explain the initial trace result. Using multiple TB reference standards and repeated evaluations allowed us to more fully capture active disease near the limit of detection and generally demonstrated consistent associations of patient characteristics with TB across definitions.

The TB burden estimated in our trace cohort is similar or higher compared to others which have used a sputum culture reference [5–9]. While TB prevalence varies by definition, we take our study’s most conservative estimate of true TB burden to be 22%, the proportion of our originally enrolled cohort (68/311) with a positive culture within three months. At the upper end of plausible true prevalence, our composite (48%) and microbiological (41%) prevalence estimates may be inflated by clinical overdiagnosis and false-positive Ultra results, respectively. Still, our results may underestimate TB prevalence among all PWTS, because two excluded groups – those empirically treated for TB before test results returned, and those requiring hospitalization -- may have particularly high likelihood of having TB disease. In general, our prevalence estimates are high enough to support treatment for most PWTS in high-burden settings when further testing and follow up is not feasible.

We were able to identify characteristics significantly associated with TB among PWTS. Consistent with other studies [6, 8, 9], we found that trace results in patients with a recent history of TB treatment were less likely to indicate current active disease. In addition, although most participants in our cohort reported TB symptoms, being asymptomatic at baseline also conferred a roughly three-fold lower risk. PWTS identified through targeted universal TB testing strategies [14, 15] may therefore be less likely to have true TB than those identified through symptom-driven testing. Finally, we found that advanced HIV was associated with microbiologically- and culture-confirmed TB among PWTS. This finding supports current WHO guidelines’ recommendation to treat patients with HIV who have a trace result [10], not only because of their elevated risk for serious illness if TB goes untreated, but also due to a higher burden of true TB.

Our results also suggest that additional diagnostic testing after a trace result can inform disease probability estimates. A normal CXR (in those without recent TB) and a normal CRP were associated with a one-and-a-half-to three-fold lower TB risk across multiple definitions, although the absolute improvement in TB status classifications (as measured by AUC) was small. In addition, where a previous review found that evidence regarding repeat Ultra testing was lacking [16], our results indicate that a negative result on a repeat Ultra test reduces TB likelihood by at least half, depending on the reference standard (**Supplemental Table 3**). Therefore, although we were unable estimate the predictive accuracy of diagnostic results within all subgroups PWTS, our cohort-wide results suggest that a negative diagnostic result on any of several widely available diagnostic tests – CXR, CRP, and repeat Ultra – could support deferring treatment for an otherwise low-risk patient whose estimated probability of TB after the initial trace result is near a clinician’s treatment threshold (**Supplemental Table 4**).

Our study has several limitations. First, limiting enrollment to patients who had not yet initiated TB treatment may have disproportionately excluded more ill patients (especially in South Africa, where nearly 1 in 10 PWTS started treatment before they could be recruited). Therefore, our results apply primarily to patients whose TB status is sufficiently uncertain that a treating clinician would defer empiric treatment based on clinical presentation alone and await diagnostic testing. Similarly, among those enrolled, participants who were more ill were more likely to be clinically diagnosed early in follow up, limiting later opportunities to detect MTB microbiologically and potentially biasing the three-month microbiological-based prevalence estimates. In addition, there may have been differences by study site in microbiological testing sensitivity (e.g. the second sputum culture specimen was collected in the early morning in Uganda, where baseline culture-positive prevalence was higher) and in clinicians’ threshold for clinical diagnosis (particularly in the context of HIV co-infection). Notably, national guidelines in Uganda advise treatment for patients with trace results, while South African guidelines defer to clinician judgment [15, 17]. Finally, the three-month timeframe of our current analysis could be too brief to detect slowly progressing disease.

In conclusion, our study suggests that the TB prevalence among PWTS is high, but that up to half of PWTS do not have clinically or microbiologically evident TB even with three months of repeated culture, molecular testing, and clinical evaluations. While roughly half of PWTS likely warrant treatment, lower-risk characteristics, favorable diagnostic results, and low mortality rates in this observed cohort, may offer reassurance to clinicians considering treatment deferral with repeated clinical review and TB testing for select patients.

## Supporting information

Supplemental Methods and Results

## Acknowledgements

This work would not be possible without the support of the staff of Harry Gwala Regional Hospital and referring primary health care clinics, the KwaZulu-Natal Department of Health, the South African National Health Laboratory Service, the TURN-TB research team and physician consultants, and the clinical and laboratory staff at Kitebi Health Centre, Kisugu Health Centre, Kisenyi Health Centre, Kawaala Health Centre, Kiswa Health Centre, Naguru Hospital, and Mulago Hospital Wards 5&6.

## Data Availability

Data available on reasonable request. Please direct such requests to Dr. Emily A. Kendall at and Dr. Paul K. Drain at.

## Author Contributions

CV drafted the statistical analysis plan, performed the data analysis, and drafted the manuscript, with supervision from EAK. RRD contributed to the analytic approach, data curation and interpretation, and editing of the manuscript. AN, PB, and GS managed data curation. KCE and MN coordinated data acquisition in Uganda, and RD and AG conducted patient visits and data acquisition in South Africa. EAK and PKD conceptualized and supervised the study, and AS, DW, and AK all contributed to administration of the project and interpretation of results. All authors reviewed and approved the final manuscript.

