## Supplemental Methods and Results for "Prevalence and predictors of tuberculosis in adults and adolescents with sputum trace Ultra results in two high-burden clinical settings"

#### Supplemental Table 1: Outcome classification criteria by reference standard and timepoint

| **Reference standard** | **Timepoint** | **Classification** | **Criteria** |
| --- | --- | --- | --- |
| Composite | Baseline | Positive | Sputum culture with MTB or recommendation to start treatment on baseline evaluation, or TB-related death |
|  |  | Negative | Not recommended to start treatment on baseline evaluation and sputum cultures negative, missing, contaminated, or positive only for nontuberculous mycobacteria |
|  |  | Excluded | Did not complete baseline evaluation |
|  | Three months | Positive | Sputum culture with MTB or recommendation to start treatment at or before the three-month evaluation, or TB-related death |
|  |  | Negative | Not recommended to start treatment and all sputum cultures negative, missing, contaminated, or positive only for nontuberculous mycobacteria through the three-month evaluation |
|  |  | Excluded | Three-month evaluation not completed and no treatment recommendation or positive sputum culture |
| Microbiological | Baseline | Positive | Sputum culture with MTB or Xpert Ultra (> trace) from baseline evaluation |
|  |  | Negative | Sputum cultures negative or positive only for nontuberculous mycobacteria, and Xpert Ultra negative, trace, or missing from baseline evaluation |
|  |  | Excluded | Sputum cultures missing or contaminated and Xpert Ultra missing from baseline evaluation |
|  | Three months | Positive | Any sputum culture with MTB or Xpert Ultra (> trace) from any sample through the time of the three-month visit |
|  |  | Negative | All sputum cultures negative or positive only for nontuberculous mycobacteria, and all Xpert Ultra results negative, trace, or missing through the time of the three-month visit |
|  |  | Excluded | Recommended to start treatment prior to the three-month visit or three-month visit not completed, and no prior positive sputum culture or Xpert Ultra result |
| Sputum-culture | Baseline | Positive | Sputum culture with MTB from the baseline evaluation |
|  |  | Negative | All sputum cultures negative or positive only for nontuberculous mycobacteria from the baseline evaluation |
|  |  | Excluded | Sputum cultures missing or contaminated from the baseline evaluation |
|  | Three months | Positive | Sputum culture with MTB from any sample through the time of the three-month evaluation |
|  |  | Negative | At least one sputum culture negative or positive only for nontuberculous mycobacteria from the three-month visit and no sputum cultures with MTB through the time of the three-month visit |
|  |  | Excluded | Recommended to start treatment prior to the three-month visit and no sputum culture with MTB, three-month visit not completed, or all sputum cultures from the three-month visit missing or contaminated |

#### Supplemental Table 2: Missing participant outcomes by reference standard, timepoint, and reason for missingness

| **Reference standard** | **Timepoint** | **Outcome unable to be assessed due to participants already having initiated treatment** | **Outcome unable to be assessed due to missed visit and/or missing data** |
| --- | --- | --- | --- |
| Composite | Baseline | *N/A* | 0 |
| Microbiological | Baseline | *N/A* | 0 |
| Sputum culture only | Baseline | *N/A* | 0 |
| Composite | Three months | *N/A* | 10 |
| Microbiological | Three months | 60 | 11 |
| Sputum culture only | Three months | 72 | 12 |

#### Sensitivity Analysis: Regression with Imputed Missing Data

We performed a sensitivity analysis using a dataset in which participants with incomplete data were retained and missing covariates were imputed based on patterns in the empiric dataset. For this analysis, multiple imputation was performed using the default statistical methods in the ‘mice’ package (version 3.16.0): predictive mean matching for numerical data, logistic regression imputation for binary data, and polytomous regression imputation for unordered categorical variables. We then applied the regression models developed in our primary analysis to this completed dataset and generated relative risk ratios with 95% confidence intervals (Supplemental Tables 5-7).

#### Estimating TB Risk among Simulated Low-Risk Patients

We generated a simulated dataset of 5,000 individuals who all had the predefined features (HIV negative, no recent history of TB, and normal baseline CXR result). All other variables were initially set as missing. We then appended the simulated dataset to the empiric dataset and used patterns in the empiric dataset to impute values for these simulated records for all variables that were not predefined. Imputation for each variable was performed using the default statistical methods in the ‘mice’ package (version 3.16.0): predictive mean matching for numerical data, logistic regression imputation for binary data, and polytomous regression imputation for unordered categorical variables. This process generated a simulated dataset of 5,000 individuals with the specified features and a variety of other characteristics that tend to accompany those features.

We then applied our CXR-added model to this simulated dataset to predict TB risk for each individual with the designated features. We report the median and interquartile range of this predicted risk across the 5,000 simulated records.

### Supplemental Results

#### Supplemental Figure 1: Study population overview by site

*Flow diagram of patients with trace results on diagnostic evaluation, study enrollment, and visit completion, with reasons for exclusion for (a) South Africa and (b) Uganda*

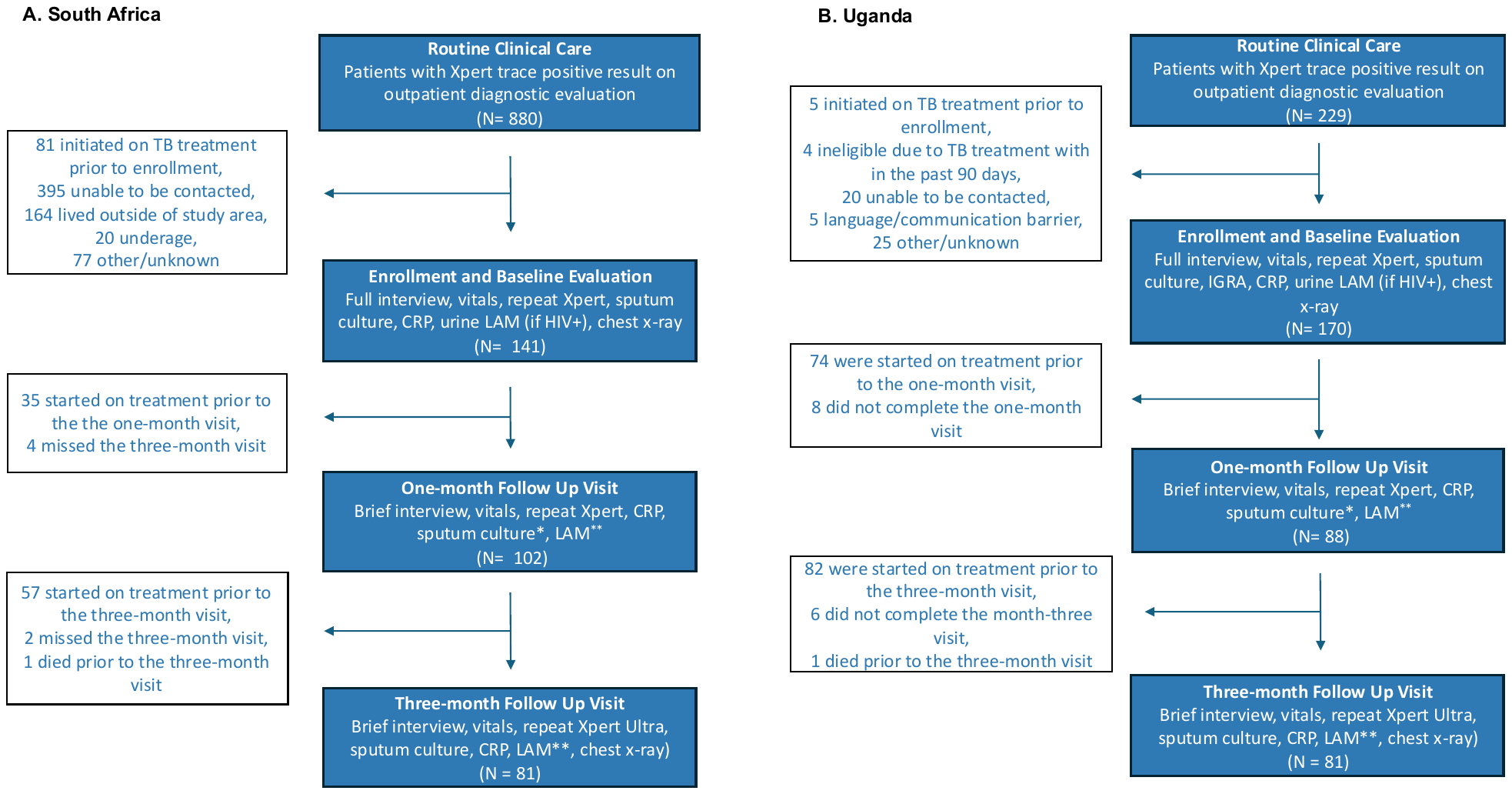

**Supplemental Figure 2: Overview of participants’ TB diagnosis status, by timing and microbiological confirmation**

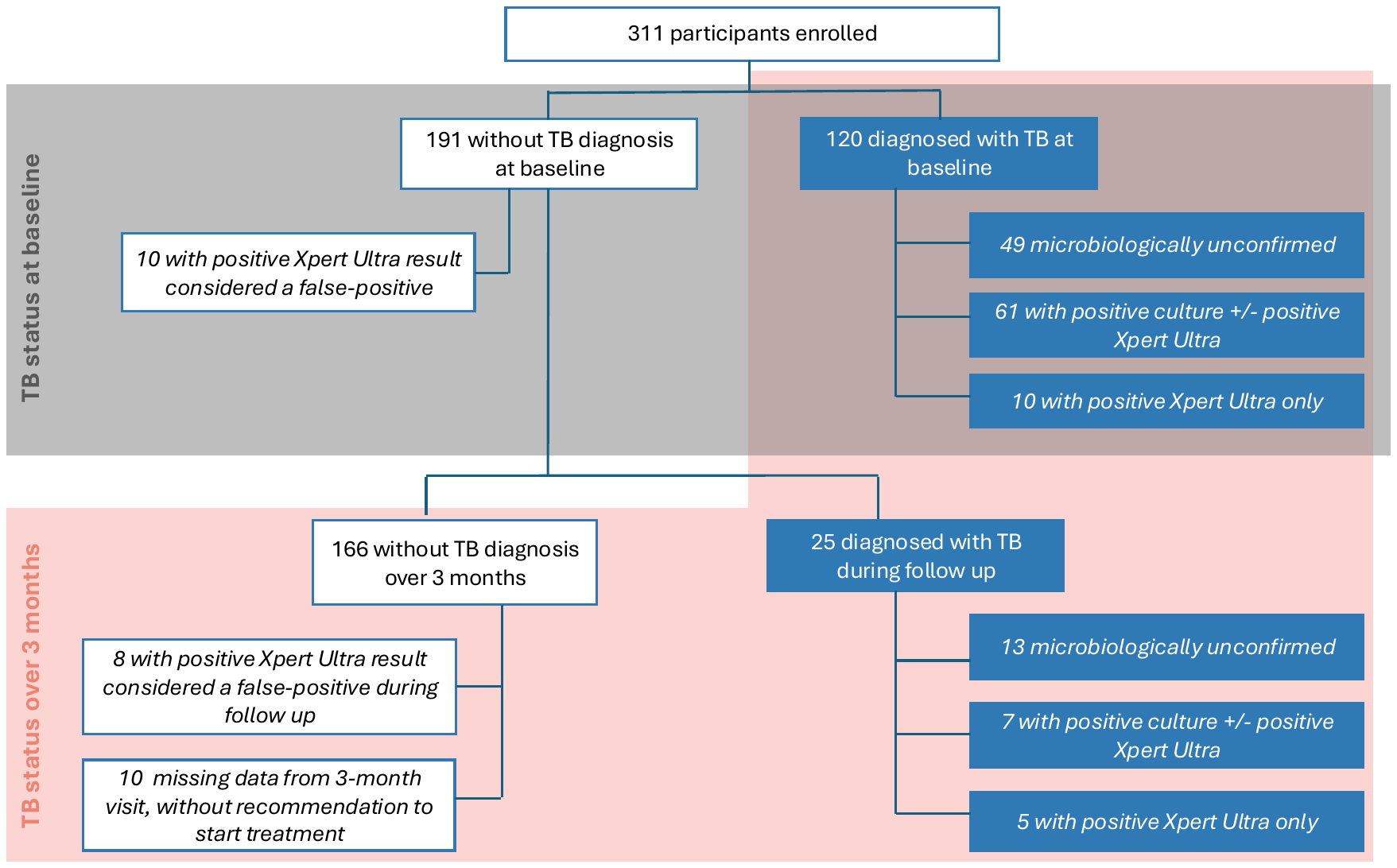

#### Supplemental Figure 3: TB prevalence among subsets of patients with initial trace Xpert Ultra results

TB prevalence by the composite, microbiological (Xpert or culture), and sputum-culture only reference standards assessed at baseline and by three months, for (A) all patients with HIV and stratified by those with CD4 counts greater than 200 cells/mm^3^and less than 200 cells/mm^3^, and (B) all patients and stratified by presence or absence of TB symptoms at enrollment. Prevalence estimates are represented as a point estimate and plotted with lines showing their 95% confidence intervals.

**A	B**

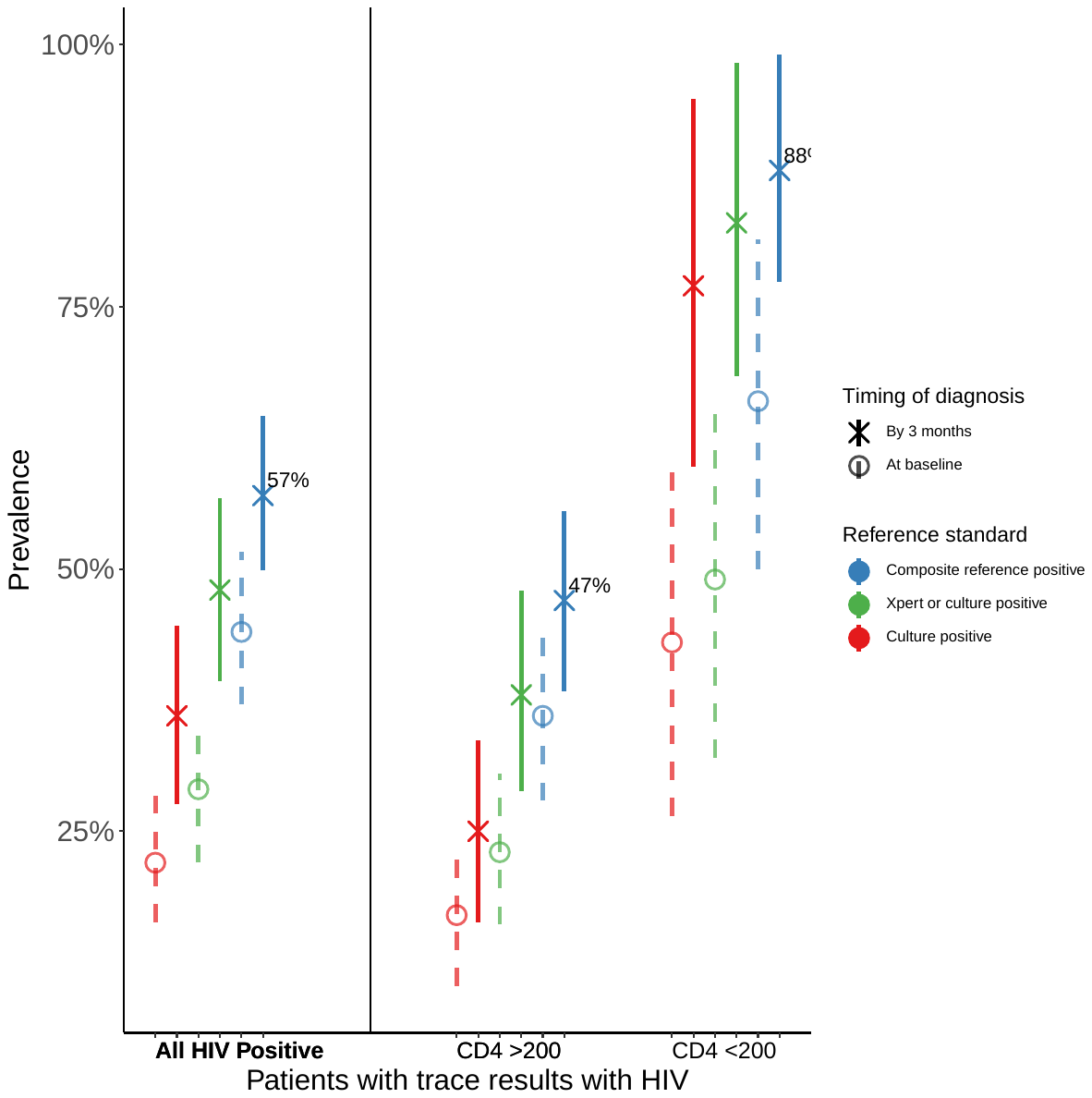

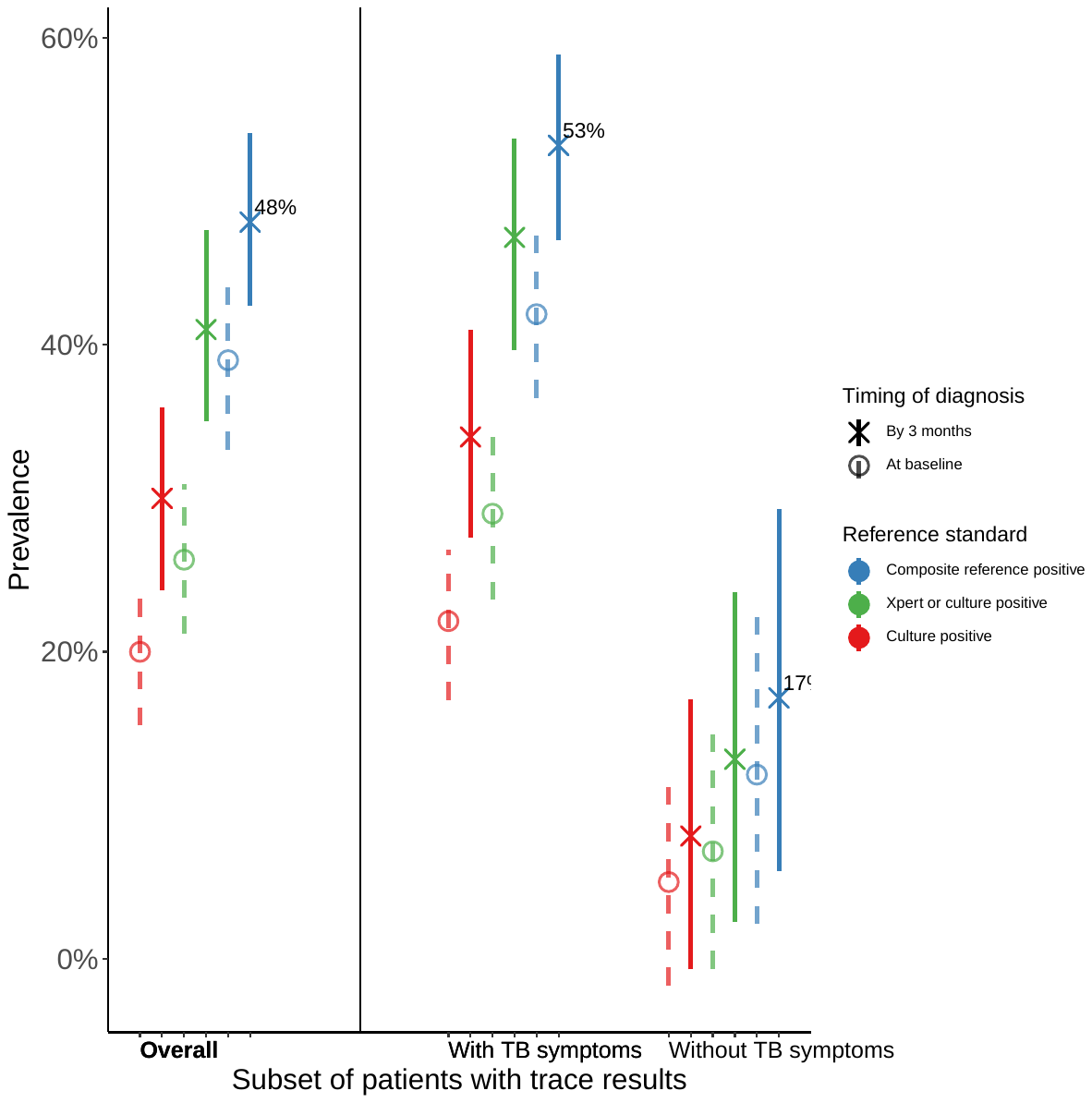

#### Supplemental Table 3: Comparison of TB prevalence ratios by patient characteristics and baseline diagnostic results among trace-positive patients using different reference standards

| **Variable** | **Prevalence ratio [95% CI]**  Composite reference standard by three months | **Prevalence ratio [95% CI]**  Microbiological reference standard by three months | **Prevalence ratio [95% CI]**  Baseline culture reference standard |
| --- | --- | --- | --- |
| Site  Uganda [reference: South Africa] | 1.2 [0.9, 1.5] | 1.0 [0.7, 1.4] | 1.7 [1.0, 2.8] |
| Age< 35 years* | 1.1 [0.9, 1.4] | 1.2 [0.9, 1.6] | 2.1 [1.3, 3.3] |
| Male sex* | 1.3 [1.0, 1.7] | 1.5 [1.1, 2.0] | 1.3 [0.8, 2.0] |
| HIV positive* | 1.6 [1.2, 2.1] | 1.4 [1.0, 2.0] | 1.4 [0.9, 2.3] |
| CD4 count <200 cells/mm^3^ (if HIV positive)* | 2.5 [1.6, 3.9] | 2.5 [1.8, 3.4] | 2.7 [1.6, 4.7] |
| Pregnant | 0.8 [0.4, 1.8] | 1.2 [0.6, 2.3] | 1.4 [0.5, 3.9] |
| Any smoking history (reference: none) | 1.1 [0.9, 1.4] | 1.3 [1.0, 1.8] | 1.1 [0.7, 1.8] |
| Current smoker (reference: never or former) | 0.9 [0.7, 1.3] | 1.0 [0.7, 1.5] | 0.6 [0.3, 1.3] |
| Positive AUDIT-C sore^1^ | 1.2 [0.9, 1.5] | 1.3 [1.0, 1.8] | 1.6 [1.0, 2.5] |
| History of TB within the past 5 years* | 0.5 [0.4, 0.8] | 0.8 [0.5, 1.2] | 0.4 [0.2, 0.8] |
| Presence of 1 or more TB symptom*^2^ | 3.0 [1.5, 6.0] | 3.5 [1.5, 8.1] | 4.4 [1.1, 17.1] |
| Recent household contact with TB* ^3^ | 0.7 [0.4, 1.2] | 0.4 [0.1, 1.0] | 0.4 [0.1, 1.5] |
| Baseline chest x-ray with any abnormality (reference: normal) | 1.7 [1.2, 2.3] | 1.7 [1.2, 2.4] | 1.3 [0.8, 2.2] |
| Baseline elevated CRP (> 5mg/L) | 2.0 [1.5, 2.6] | 2.6 [1.8, 3.7] | 4.0 [2.2, 7.2] |
| Baseline repeat Xpert Ultra positive or trace (reference: negative) | 2.2 [1.8, 2.7] | 4.4 [2.9, 6.8] | 5.9 [3.6, 9.6] |

*Variables included in the multivariate regression models shown in Table 2, Supplemental Table 5, and Supplemental Table 6

AUDIT-C = Alcohol Use Disorders Identification Test

1. Positive AUDIT-C score defined as 4 or greater for men and 3 or greater for women.

2. Defined as cough, fever, weight loss, or night sweats at time of study enrollment

3. Defined as living with someone diagnosed with TB within the past 6 months

#### Supplemental Table 4: Frequency of baseline diagnostic testing results among trace-positive participants by TB outcome, by multiple TB reference standards

Frequency of each diagnostic testing report reported for participants according to whether they were classified as positive or negative for each of the following reference standards: composite reference standard, assessed at three months; microbiological (Xpert or culture) reference standard, assessed at three months; sputum culture reference standard, assessed at baseline. Participants with missing results were excluded from the denominators.

|  | **Reference standard categorization** | | | | | |
| --- | --- | --- | --- | --- | --- | --- |
|  | **Composite reference standard by three months** | | **Microbiological reference standard by three months** | | **Baseline sputum culture reference standard** | |
| **Diagnostic testing result** | **Positive**  **(N=145)** | **Negative (N=156)** | **Positive**  **(N=99)** | **Negative**  **(N=141)** | **Positive (N=61)** | **Negative**  **(N=250)** |
| Baseline CXR abnormal (all) (n/N, [%])^[1]^ | 108/142 (76%) | 84/152 (55%) | 70/97  (72%) | 72/136  (53%) | 42/59  (71%) | 154/24  (63%) |
| Baseline CXR abnormal (no recent TB history) (n/N, [%]) | 87/118^2^  (74%) | 39/106^3^ (37%) | 53/77^4^  (69%) | 36/97^5^  (37%) | 37/53^6^  (70%) | 91/174^7^  (52%) |
| Baseline elevated CRP (>=5 mg/L) (n/N, [%])^8^ | 96/144  (67%) | 54/156  (35%) | 70/98  (71%) | 47/141  (33%) | 48/60  (80%) | 107/250  (43%) |
| Repeat baseline Xpert positive (>trace) (n/N, [%])^9^ | 44/142  (31%) | 8/150  (5%) | 52/96  (54%) | 0/137  (0%) | 32/61  (52%) | 20/241  (8%) |
| Repeat baseline Xpert trace (n/N, [%])^9^ | 24/142  (17%) | 10/150  (7%) | 13/96  (14%) | 10/137  (7%) | 11/61  (18%) | 24/241  (10%) |
| Repeat baseline Xpert negative (n/N, [%])^9^ | 74/142  (52%) | 132/150  (88%) | 31/96  (32%) | 127/137  (93%) | 18/61  (30%) | 197/241  (82%) |

1. Differences between cell N and column N represent the number of participants with missing baseline CXR results.
2. 120 participants had no recent TB history, and 2 of these were missing baseline CXR results and excluded.
3. 106 participants had no recent TB history.
4. 78 participants had no recent TB history, and 1 of these was missing a baseline CXR result and excluded.
5. 101 participants had no recent TB history, and 4 of these were missing a baseline CXR result and excluded.
6. 54 participants had no recent TB history, and 1 of these was missing a baseline CXR result and excluded.
7. 179 participants had no recent TB history, and 5 of these were missing baseline CXR results and excluded.
8. Differences between cell N and column N represent the number of participants with missing baseline CRP results.
9. Differences between cell N and column N represent the number of participants with missing baseline repeat Xpert results.

#### Supplemental Table 5: Multivariate log-binomial regression model coefficients for discriminating TB vs no TB, using microbiologically positive reference standard over 3 months

| **Covariate** | **Unadjusted prevalence ratio (95% CI), n=230^3^** | **Adjusted prevalence ratio (95% CI) for variables included in final model** | | |
| --- | --- | --- | --- | --- |
|  |  | **Patient characteristics only, n=238^4^** | **Addition of CXR, n=231^5^** | **Addition of CRP, n=237^6^** |
| Uganda site | 1.0 (0.7, 1.4) | - | - | - |
| Age <35 | 1.2 (0.9, 1.7) | 1.2 (0.8, 1.8) | 1.3 (0.8, 1.9) | 1.2 (0.8, 1.9) |
| Male gender | 1.5 (1.1, 2.0) | 1.4 (0.9, 2.1) | 1.3 (0.8, 2.0) | 1.3 (0.8, 2.0) |
| History of prior TB within 5 years | 0.8 (0.5, 1.1) | 0.8 (0.5, 1.3) | 1.3 (0.3, 3.9) | 0.8 (0.5, 1.4) |
| Presence of 1 or more TB symptom^1^ | 3.3 (1.7, 9.0) | 3 (1.3, 8.7) | 2.6 (1.1, 7.4) | 2.6 (1.2, 7.6) |
| Recent household TB contact^2^ | 0.4 (0.1, 0.8) | 0.4 (0.1, 1.1) | 0.5 (0.1, 1.3) | 0.4 (0.1, 1.2) |
| Any smoking history | 1.3 (1.0, 1.8) | - | - | - |
| HIV positive (all)  HIV positive with CD4 <200 cells/mm^3^  HIV positive with CD4 >200 cells/mm^3^  HIV positive with CD4 unknown | 1.5 (1.1, 2.0)  2.5 (1.4, 4.3)  1.2 (0.7, 1.9)  2.2 (0.5, 6.2) | -  2.1 (1.2, 3.6)  1.3 (0.8, 2.0)  1.9 (0.5, 5.3) | -  2.0 (1.1, 3.5)  1.3 (0.8, 2.1)  2.1 (0.5, 6.0) | -  1.7 (0.9, 2.9)  1.4 (0.8, 2.2)  2.1 (0.5, 6.0) |
| Abnormal CXR (overall)  Abnormal CXR in those without history of TB within 5 years  Abnormal CXR in those with history of TB within 5 years | 1.6 (1.2, 2.4)  2.1 (1.3, 3.5)    0.3 (0.1, 1.4) | *-*  -  - | -  1.6 (1.0, 2.7)  0.5 (0.1, 2.3) | *-*  -  - |
| CRP > 5mg/L | 2.5 (1.8, 3.7) | *-* | - | 2.1 (1.3, 3.3) |

1. Defined as cough, fever, weight loss, or night sweats at time of study enrollment

2. Defined as living with someone diagnosed with TB within the past 6 months

3. 81 observations dropped due to incomplete data

4. 73 observations dropped due to incomplete data

5. 80 observations dropped due to incomplete data

6. 74 observations dropped due to incomplete data

#### Supplemental Table 6: Multivariate log-binomial regression model coefficients for discriminating TB vs no TB, using culture positive at baseline reference standard

| **Covariate** | **Unadjusted prevalence ratio (95% CI), n=295^3^** | **Adjusted prevalence ratio (95% CI) for variables included in final model** | | |
| --- | --- | --- | --- | --- |
|  |  | **Patient characteristics only, n=305^4^** | **Addition of CXR, n=296^5^** | **Addition of CRP, n=304^6^** |
| Uganda site | 1.6 (1.01, 2.8) | - | - | - |
| Age < 35 | 2.19 (1.37, 3.57) | 2.1 (1.3, 3.6) | 2.2 (1.3, 3.8) | 2.1 (1.2, 3.5) |
| Male gender | 1.2 (0.8, 2) | 1.2 (0.7, 2.1) | 1.2 (0.7, 2.0) | 1.1 (0.7, 2.0) |
| History of prior TB within 5 years | 0.3 (0.1, 0.7) | 0.4 (0.2, 0.9) | 0.8 (0.0, 3.9) | 0.4 (0.2, 0.9) |
| Presence of 1 or more TB symptom^1^ | 3.9 (1.3, 23.5) | 3.7 (1.1, 23.0) | 3.1 (0.9, 19.1) | 2.7 (0.8, 17.0) |
| Recent household TB contact^2^ | 0.4 (0.1, 1.2) | 0.4 (0.1, 1.3) | 0.4 (0.1, 1.4) | 0.4 (0.1, 1.4) |
| Any smoking history | 1.1 (0.7, 1.8) | - | - | - |
| HIV positive (all)  HIV positive with CD4 <200 cells/mm^3^  HIV positive with CD4 >200 cells/mm^3^  HIV positive with CD4 unknown | 1.5 (0.9, 2.5)  2.9 (1.6, 5.1)  1.1 (0.6, 2.0)  2.0 (0.5, 4.7) | -  2.3 (1.1, 4.6)  1.1 (0.6, 2.1)  1.3 (0.3, 4.0) | -  2.2 (1.1, 4.5)  1.2 (0.6, 2.2)  1.5 (0.3, 4.6) | -  1.8 (0.9, 3.6)  1.3 (0.7, 2.4)  1.5 (0.4, 4.5) |
| Abnormal CXR (overall)  Abnormal CXR in those without history of TB within 5 years  Abnormal CXR in those with history of TB within 5 years | 1.3 (0.8, 2.2)  1.8 (1.1, 3.1)      0.2 (0.0, 4.0) | -  -  - | -  1.4 (0.8, 2.8)      0.4 (0.0, 7.9) | *-*  -  - |
| CRP > 5mg/L | 3.8 (2.2, 7.2) | *-* | - | 3.2 (1.7, 6.4) |

1. Defined as cough, fever, weight loss, or night sweats at time of study enrollment

2. Defined as living with someone diagnosed with TB within the past 6 months

3. 16 observations dropped due to incomplete data

4. 6 observations dropped due to incomplete data

5. 15 observations dropped due to incomplete data

6. 7 observations dropped due to incomplete data

#### Supplemental Table 7: Multivariate log-binomial regression model coefficients for discriminating TB vs no TB, using composite reference standard over 3 months, with missing data completed by multiple imputation

| **Covariate** | **Unadjusted prevalence ratio (95% CI), n=311** | **Adjusted prevalence ratio (95% CI) for variables included in final model** | | |
| --- | --- | --- | --- | --- |
|  |  | **Patient characteristics model, n=311** | **CXR-added model, n=311** | **CRP-added model, n=311** |
| Uganda site | 1.1 (0.9, 1.4) | - | - | - |
| Age < 35 | 1.1 (0.9, 1.4) | 1.1 (0.8, 1.6) | 1.2 (0.8, 1.6) | 1.1 (0.8, 1.6) |
| Male gender | 1.2 (1.0, 1.6) | 1.2 (0.9, 1.7) | 1.1 (0.8, 1.6) | 1.2 (0.8, 1.7) |
| History of prior TB within 5 years | 0.5 (0.3, 0.8) | 0.6 (0.3, 0.9) | 0.8 (0.1, 2.8) | 0.6 (0.4, 0.9) |
| Presence of 1 or more TB symptom^1^ | 3.0 (1.7, 6.7) | 2.7 (1.3, 6.4) | 2.3 (1.1, 5.5) | 2.4 (1.2, 5.8) |
| Recent household TB contact^2^ | 0.7 (0.4, 1.1) | 0.8 (0.4, 1.5) | 0.9 (0.4, 1.6) | 0.8 (0.4, 1.5) |
| Any smoking history | 1.1 (0.9, 1.4) | - | - | - |
| HIV positive (all)  HIV positive with CD4 <200 cells/mm^3^  HIV positive with CD4 >200 cells/mm^3^  HIV positive with CD4 unknown | 1.6 (1.3, 2.2)  2.5 (1.6, 3.9)  1.3 (0.9, 2.0)  2.5 (1.2, 4.9) | -  2.1 (1.3, 3.3)  1.4 (0.9, 2.1)  2.1 (0.9, 4.1) | -  2.0 (1.3, 3.2)  1.4 (1.0, 2.1)  2.4 (1.1, 4.7) | -  1.8 (1.1, 2.9)  1.5 (1, 2.2)  2.1 (1, 4.2) |
| Abnormal CXR (overall)  Abnormal CXR in those without history of TB within 5 years  Abnormal CXR in those with history of TB within 5 years | 1.6 (1.2, 2.2)  2.0 (1.4, 3.0)  0.4 (0.1, 2.7) | *-*  -  - | -  1.8 (1.2, 2.7)    0.6 (0.2, 3.7) | *-*  -  - |
| CRP > 5mg/L | 2.0 (1.6, 2.6) | *-* | - | 1.7 (1.2, 2.4) |

1. Defined as cough, fever, weight loss, or night sweats at time of study enrollment

2. Defined as living with someone diagnosed with TB within the past 6 months

#### Supplemental Figure 4: Receiver operating characteristic curves for the basic characteristics, CXR-added, and CRP-added models, using (A) the microbiological reference standard over three months and (B) the baseline culture reference standard

**A	B**

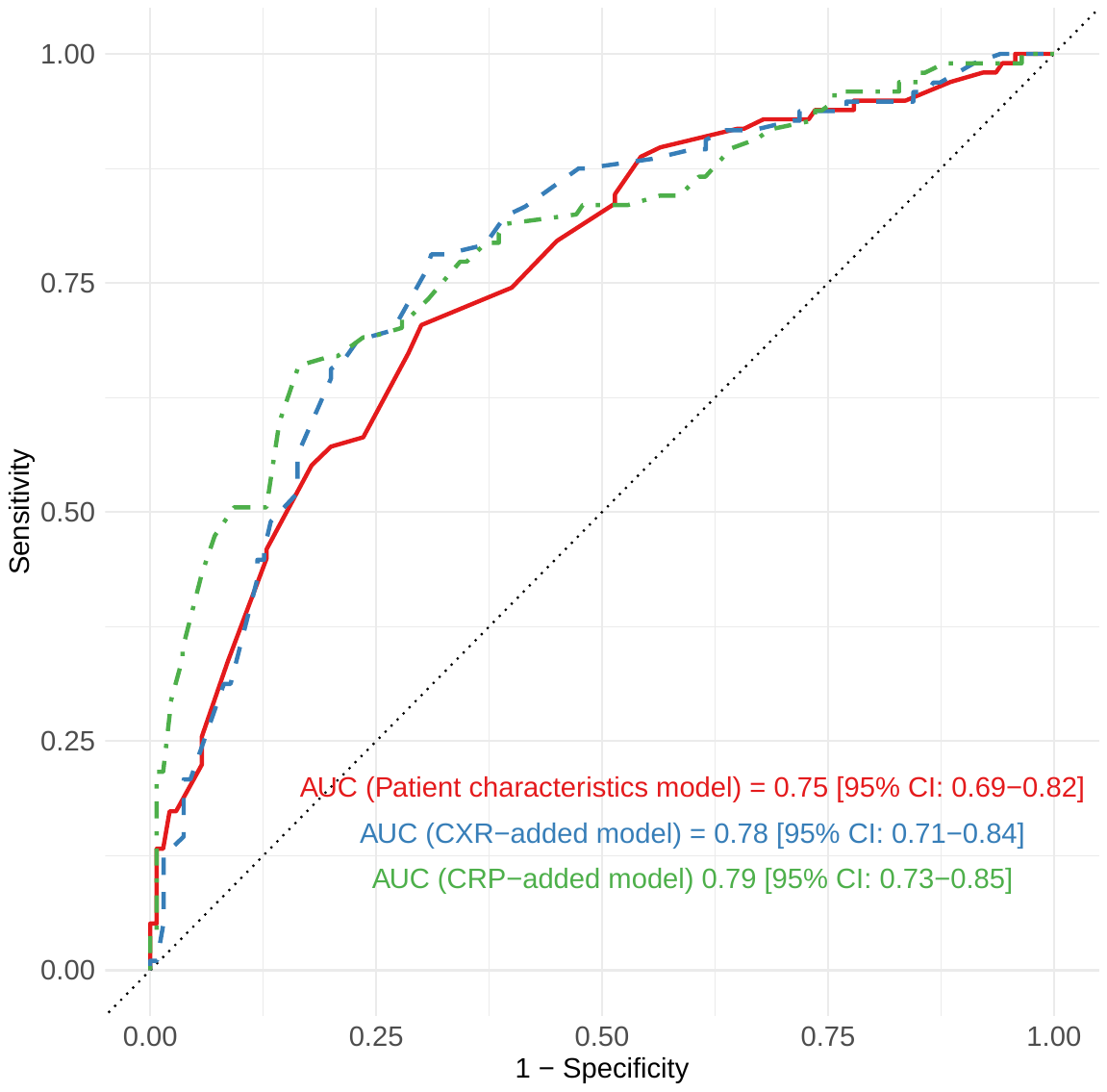

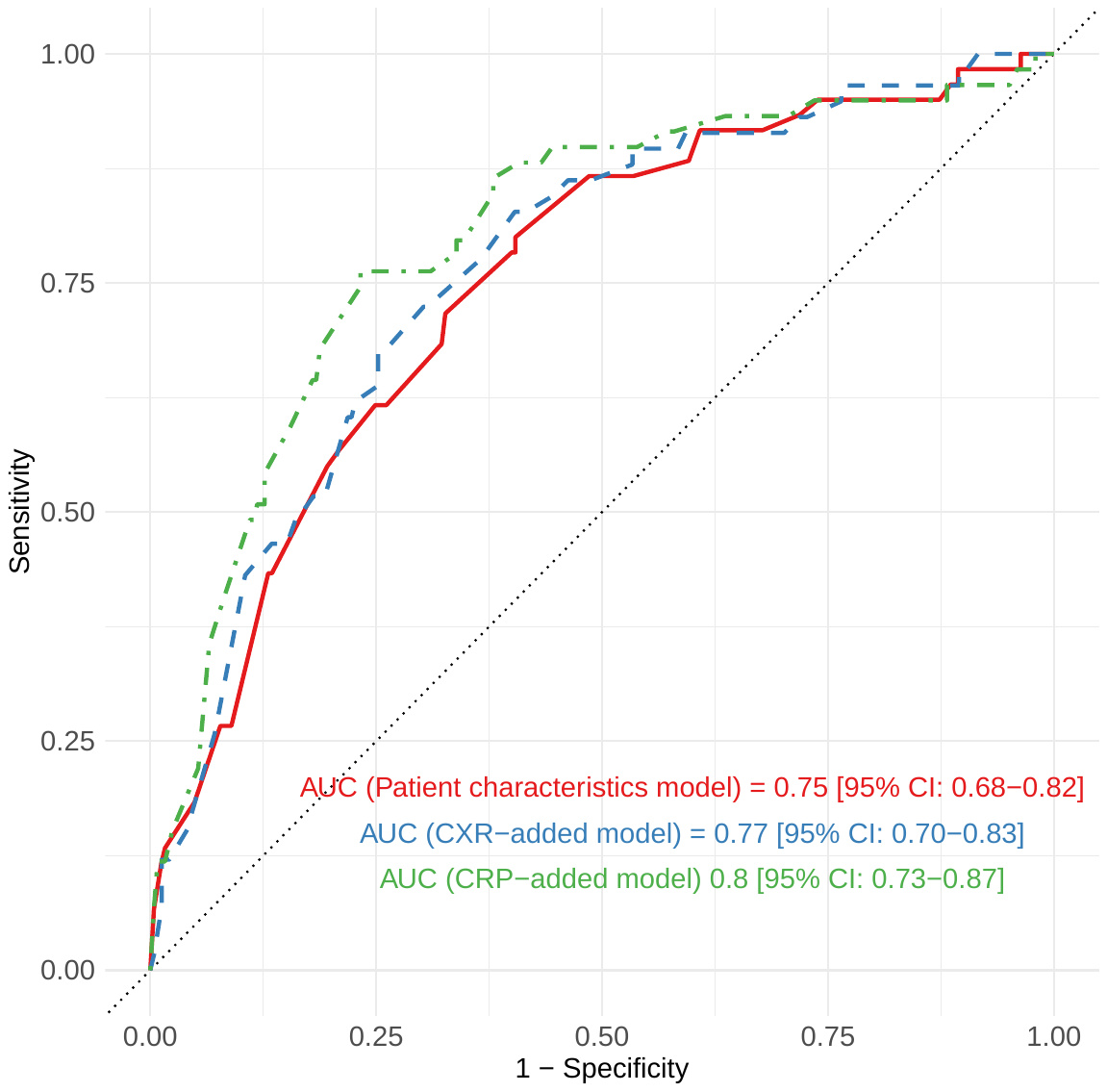

*AUC = Area-under-the-curve*

*CI = Confidence interval*
